# Impact of Multiple Episodes of Cytomegalovirus Infection on Patient Outcomes in Allogeneic Hematopoietic Stem Cell Transplant Using a Pre-emptive Monitoring Strategy

**DOI:** 10.1101/2024.08.27.24312676

**Authors:** Julia A. Messina, Jennifer L. Saullo, Steven Wolf, Yanhong Li, Helen Tang, Lauren B. Hill, Gena G. Foster, Michael Grant, Samantha M. Thomas, Rachel A. Miller, Barbara D. Alexander, Mitchell Horwitz, Anthony D. Sung

**Author notes:** **Corresponding author:** Julia A. Messina, MD, MHS, MS 315 Trent Drive, Durham, NC, USA, 27705.

## Abstract

The impact of multiple episodes of clinically significant cytomegalovirus infection (csCMVi) on clinical outcomes during pre-emptive CMV monitoring in allogeneic hematopoietic stem cell transplant recipients (HCT) is not well understood.

We performed a retrospective cohort study of all consecutive patients undergoing their first allogeneic HCT at Duke between January 1, 2009 and December 31, 2013, during an era of pre-emptive CMV monitoring, prior to widespread use of letermovir prophylaxis. Consensus definitions were utilized for refractory and resistant CMV infection, bacteremia, invasive fungal disease, and renal dysfunction. The Kaplan-Meier method was used to estimate survival.

Three hundred eighty-eight adult patients underwent allogeneic HCT during the study time period with 91 patients (23%) having one episode of csCMVi and 79 patients (20%) having ≥ one episode of csCMVi. Risk factors for multiple episodes of csCMVi included having a transplant from a matched unrelated donor, cord blood as HCT source, and graft-versus-host disease prophylaxis with highly T-cell suppressive agents such as alemtuzumab. Patients with multiple episodes tended to develop csCMVi earlier post-transplant and were more likely to have CMV disease and to develop resistant CMV infection. There was no difference in rates of refractory CMV infection, bacteremia, invasive fungal disease, or survival. However, patients with multiple episodes of csCMV had reduced one-year relapse-related mortality compared to patients with one csCMVi episode.

## Introduction

Cytomegalovirus (CMV) infection remains a critical cause of morbidity and mortality in the allogeneic hematopoietic cell transplant (HCT) population despite improvements in the management of CMV infection. Prior studies have shown that CMV infection increases the risk for invasive fungal disease (IFD), graft-versus-host disease (GVHD), and non-relapse related mortality (NRM) [1–3]. We previously reported that clinically significant CMV infection (csCMVi), defined as CMV DNAemia necessitating preemptive therapy or CMV disease, is also significantly associated with increased total costs and hospital length-of-stay [4].

An important unanswered question is the impact of multiple episodes of csCMVi on patient outcomes, which we seek to answer in the current study performed during an era of pre-emptive CMV monitoring. We hypothesize that patients with multiple episodes of csCMVi have more frequent resistant and refractory infection and CMV disease as defined by consensus definitions devised by expert panels [5, 6]. We also hypothesize that multiple episodes of csCMVi are associated with increased renal dysfunction due to use of nephrotoxic drugs such as foscarnet as salvage therapy for refractory or resistant infection. A final objective is to investigate the impact of multiple episodes of csCMVi on clinical outcomes including invasive bacterial infections, IFD, disease relapse, survival, and renal function through day +365 post-transplant.

## Methods and methods

### Study patients

This retrospective cohort study received approval by the Duke Health Institutional Review Board, and a waiver of informed consent was obtained. All consecutive patients undergoing their first allogeneic HCT at Duke University Medical Center between January 1, 2009 and December 31, 2013 were reviewed. We specifically chose a study period that preceded the Food and Drug Administration approval of letermovir in 2017 for universal prophylaxis of high-risk CMV HCT recipients. During the study period, all patients underwent CMV pre-emptive monitoring regardless of CMV serostatus.

The following were excluded from the analysis: patients receiving the investigational agent brincidofovir post-transplant (N=8), with syngeneic donors (N=3), having CMV infection and/or disease within two months preceding allogeneic HCT (N=3), or undergoing sequential organ transplant followed by HCT (N=1, lung-HCT transplant). This study was approved by the Institutional Review Board of Duke University Health System (DUHS). Demographic and clinical data were abstracted from a prospectively maintained Duke Adult Blood and Marrow Transplant database, an institutional tool called Duke Enterprise Data Unified Content Explorer, and manual review of electronic medical records.

### Antimicrobial prophylaxis

Patients were pre-emptively monitored via plasma quantitative CMV polymerase chain reaction (PCR) using a DUHS laboratory-developed test incorporating the *artus* CMV PCR kit (Qiagen, Hilden, Germany) for amplification of a specific region of the CMV genome. Extraction of CMV DNA was performed on the MagNA Pure Compact (Roche), and amplification and detection were performed on the Lightcycler (Roche, Indianapolis, IN). Plasma CMV PCR evaluation occurred at least weekly beginning the first week post-transplant through a minimum of day +100. Monitoring beyond day +100 was continued at providers’ discretion in patients receiving ongoing immunosuppression. CMV-directed antiviral therapy (e.g., ganciclovir, foscarnet, or valganciclovir) was initiated in patients with 1) evidence of or concern for CMV disease; or 2) when the plasma CMV DNA PCR exceeded a designated threshold value. The recommended threshold value was greater than 250 copies/milliliter (mL) through April 2014 with modification thereafter to greater than 450 international units (IU)/mL following an update to the plasma CMV DNA PCR reporting. Per institutional practice, the minimum suggested duration, for induction therapy was two weeks followed by an additional two weeks of maintenance therapy. Patients not receiving CMV-directed therapies were maintained on herpes virus prophylaxis with acyclovir. Recommended bacterial prophylaxis was ciprofloxacin through a minimum period of engraftment. Fungal prophylactic regimens included either fluconazole, voriconazole, or posaconazole through at least day +100. *Pneumocystis jirovecii* (PJP) prophylaxis consisted of sulfamethoxazole/trimethoprim during conditioning through day -2 with resumption following engraftment or day +30.

### Clinical definitions

Clinically significant CMV infection (csCMVi) was defined as CMV DNAemia necessitating preemptive therapy or CMV disease [7]. Assessment for CMV disease was based on standardized definitions [5, 6]. Delineation of a second or subsequent episode of csCMVi required a minimum of a two-week period following completion of CMV-directed therapy. CMV viral resistance was determined via a CMV genotype demonstrating the presence of a UL97 or UL54 mutation confirmed in previous studies to confer antiviral resistance through marker transfer experiments [8]. Resistant CMV infection was defined as the presence of viral genetic alteration that decreases susceptibility to one or more antiviral drugs.[5] Refractory csCMVi was defined as greater than one log_10_ increase in CMV DNA levels in blood or plasma and determined by log_10_ change from the peak viral load within the first week to the peak viral load at greater than or equal to two weeks as measured in the same laboratory with the same assay [5]. Probable refractory csCMVi was defined as persistent viral load (same or higher than peak viral load within one week but less than one log_10_ increase) after at least two weeks of appropriately dosed antiviral therapy as measured in the same laboratory with the same assay. Refractory CMV end-organ disease was defined as worsening signs and/or symptoms or progression to end-organ disease after at least two weeks of appropriate treatment. Probable refractory end-organ disease was defined as lack of improvement in signs and symptoms after at least two weeks of appropriately dosed antiviral drugs.

Bacteremia was defined as a recognized pathogen from one or more blood cultures. More than one episode of bacteremia was recorded in the same patient only if it occurred after a minimum of two weeks from the previous positive blood culture in patients receiving directed therapy. Common skin commensals, as defined by the National Healthcare Safety Network, were designated causes of bacteremia only if isolated from two or more blood specimens drawn on separate occasions meeting criteria that blood from at least two separate blood draws was collected on the same or consecutive calendar days, and from two separate blood draw sites [9]. Proven and probable invasive fungal infections were based on modified criteria proposed by the European Organization for Research and Treatment of Cancer/ Mycoses Study Group [10]. These definitions were further modified to allow for inclusion of PCR testing from bronchoalveolar lavage specimens to identify probable PJP pulmonary infections. Acute and chronic GVHD were scored based on standardized criteria [11–13].

Definitions for renal dysfunction were included in a previous publication on foscarnet use in allogeneic HCT recipients [14]. Estimated glomerular filtration rate (eGFR) was calculated based on the Chronic Kidney Disease Epidemiology Collaboration (CKD-EPI) formula [15]. Acute kidney injury (AKI) and acute kidney failure was defined as a creatinine increase of at least two and three times the baseline Cr, or >50% and >75% decrease in eGFR, respectively [16]. eGFR at three months was calculated using the median of all creatinine measures taken within 15 days of day +90, eGFR at six months was calculated using the median of all creatinine measures taken within 45 days of day +180, and eGFR at 12 months was defined as the median of all measures taken within 45 days of day +365. Comparisons in renal dysfunction were made between patients without csCMVi, those with one episode of csCMVi, and those with multiple episodes of csCMVi.

Overall survival (OS) was defined as time to death from any cause or last follow up from the day of transplant. Non-relapse related mortality (NRM) was defined as death without evidence of disease relapse. Patients who did not experience events were censored on the date of last follow-up. Disease relapse was defined as the recurrence of the underlying hematologic malignancy after HCT while relapse-related mortality (RRM) was defined as death with evidence of disease relapse.

### Statistical analyses

Descriptive statistics were used to report baseline demographics, clinical characteristics, and outcomes. Percentages and numbers were reported for categorical variables. Means, standard deviations, medians, and the 25^th^ and 75^th^ percentiles were reported for continuous variables. In comparisons of baseline characteristics between patient groups, Chi-squared or Fisher’s exact tests were used for categorical variables, and Wilcoxon rank-sum tests were used for continuous variables. We examined change in renal function within patients from baseline to 90 days, 6 months and 12 months, we compared patients with at least one csCMVi to patients who didn’t have any csCMVi events. We tested for differences between csCMVi patients to those who didn’t have csCMVi using the Wilcoxon rank-sum test for continuous variables and either a Chi-sqaured or Fisher’s exact test where appropriate.

The Kaplan-Meier method was used to estimate overall survival, and NRM and RRM as competing-risk outcomes. Comparisons between patient groups were based on log-rank tests. We compared patients based on whether they had one csCMVi episode, more than one csCMVi episode, or no csCMVi episodes. All statistical analyses were conducted using SAS Version 9.4 (SAS Institute, Cary, NC).

## Results and discussion

During the study period, 388 adult patients underwent allogeneic HCT at Duke with 170 patients (43.8%) having at least one episode of csCMVi (Table 1). Among the 170 patients with csCMVi, 91 patients (54%) had one episode of csCMVi while 50 patients (29%) had two episodes, 15 patients (9%) had three episodes, 12 patients (7%) had four episodes, and two patients (1%) had five episodes. Thirty-seven (21.8%) patients had CMV disease, and the most common site of CMV disease was the gastrointestinal tract (28/37 patients; 75.7%) (Table 2).

**Table 1:**
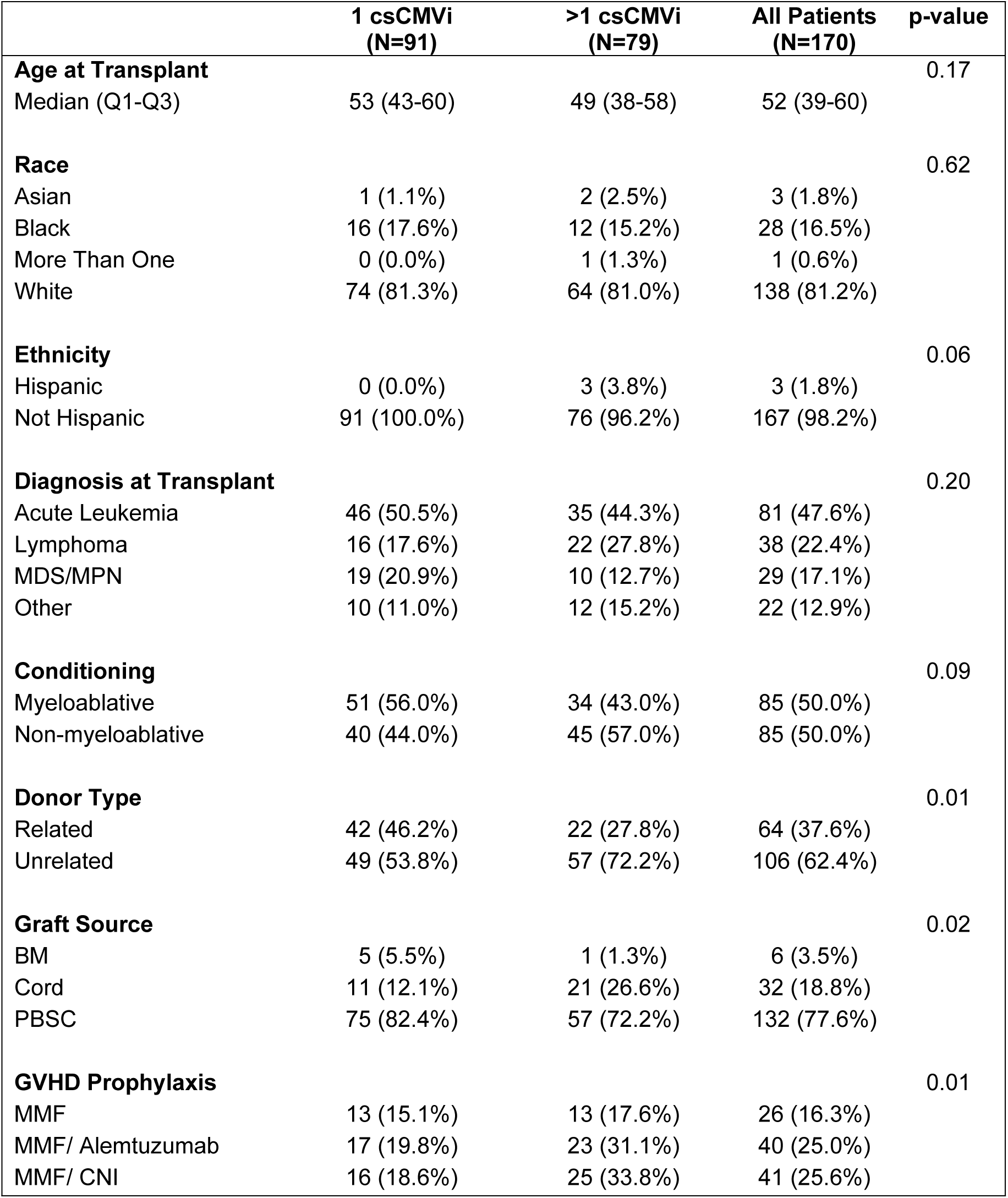
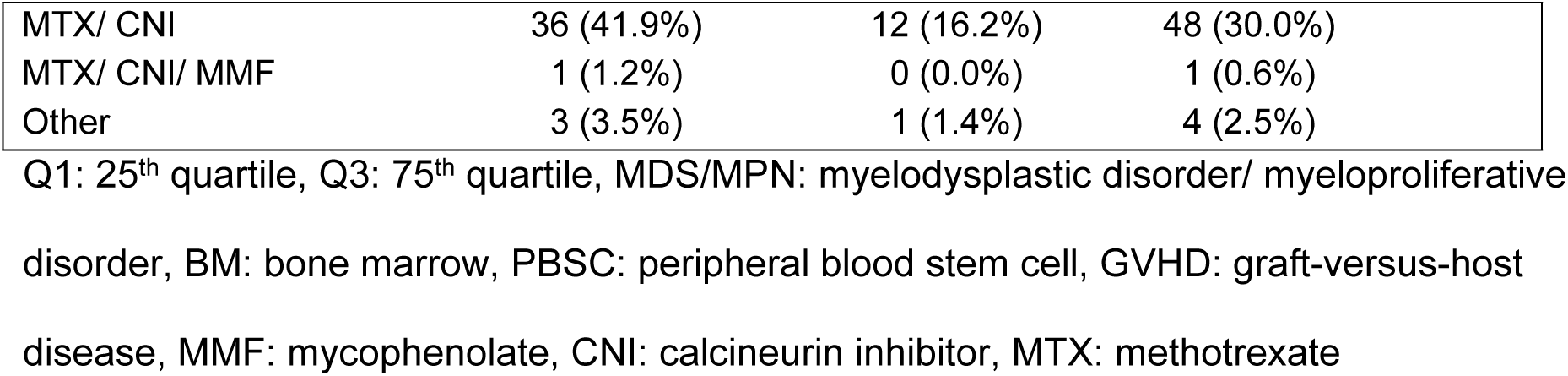
Baseline Patient Demographics.

|  | **1 csCMVi**<br>**(N=91)** | **>1 csCMVi**<br>**(N=79)** | **All Patients**<br>**(N=170)** | **p-value** |
| --- | --- | --- | --- | --- |
| **Age at Transplant** |  |  |  | 0.17 |
| Median (Q1-Q3) | 53 (43-60) | 49 (38-58) | 52 (39-60) |  |
| **Race** |  |  |  | 0.62 |
| Asian | 1 (1.1%) | 2 (2.5%) | 3 (1.8%) |  |
| Black | 16 (17.6%) | 12 (15.2%) | 28 (16.5%) |  |
| More Than One | 0 (0.0%) | 1 (1.3%) | 1 (0.6%) |  |
| White | 74 (81.3%) | 64 (81.0%) | 138 (81.2%) |  |
| **Ethnicity** |  |  |  | 0.06 |
| Hispanic | 0 (0.0%) | 3 (3.8%) | 3 (1.8%) |  |
| Not Hispanic | 91 (100.0%) | 76 (96.2%) | 167 (98.2%) |  |
| **Diagnosis at Transplant** |  |  |  | 0.20 |
| Acute Leukemia | 46 (50.5%) | 35 (44.3%) | 81 (47.6%) |  |
| Lymphoma | 16 (17.6%) | 22 (27.8%) | 38 (22.4%) |  |
| MDS/MPN | 19 (20.9%) | 10 (12.7%) | 29 (17.1%) |  |
| Other | 10 (11.0%) | 12 (15.2%) | 22 (12.9%) |  |
| **Conditioning** |  |  |  | 0.09 |
| Myeloablative | 51 (56.0%) | 34 (43.0%) | 85 (50.0%) |  |
| Non-myeloablative | 40 (44.0%) | 45 (57.0%) | 85 (50.0%) |  |
| **Donor Type** |  |  |  | 0.01 |
| Related | 42 (46.2%) | 22 (27.8%) | 64 (37.6%) |  |
| Unrelated | 49 (53.8%) | 57 (72.2%) | 106 (62.4%) |  |
| **Graft Source** |  |  |  | 0.02 |
| BM | 5 (5.5%) | 1 (1.3%) | 6 (3.5%) |  |
| Cord | 11 (12.1%) | 21 (26.6%) | 32 (18.8%) |  |
| PBSC | 75 (82.4%) | 57 (72.2%) | 132 (77.6%) |  |
| **GVHD Prophylaxis** |  |  |  | 0.01 |
| MMF | 13 (15.1%) | 13 (17.6%) | 26 (16.3%) |  |
| MMF/ Alemtuzumab | 17 (19.8%) | 23 (31.1%) | 40 (25.0%) |  |
| MMF/ CNI | 16 (18.6%) | 25 (33.8%) | 41 (25.6%) |  |

|  |  |  |  |
| --- | --- | --- | --- |
| MTX/ CNI | 36 (41.9%) | 12 (16.2%) | 48 (30.0%) |
| MTX/ CNI/ MMF | 1 (1.2%) | 0 (0.0%) | 1 (0.6%) |
| Other | 3 (3.5%) | 1 (1.4%) | 4 (2.5%) |

Q1: 25<sup>th</sup> quartile, Q3: 75<sup>th</sup> quartile, MDS/MPN: myelodysplastic disorder/ myeloproliferative disorder, BM: bone marrow, PBSC: peripheral blood stem cell, GVHD: graft-versus-host disease, MMF: mycophenolate, CNI: calcineurin inhibitor, MTX: methotrexate

**Table 2:**
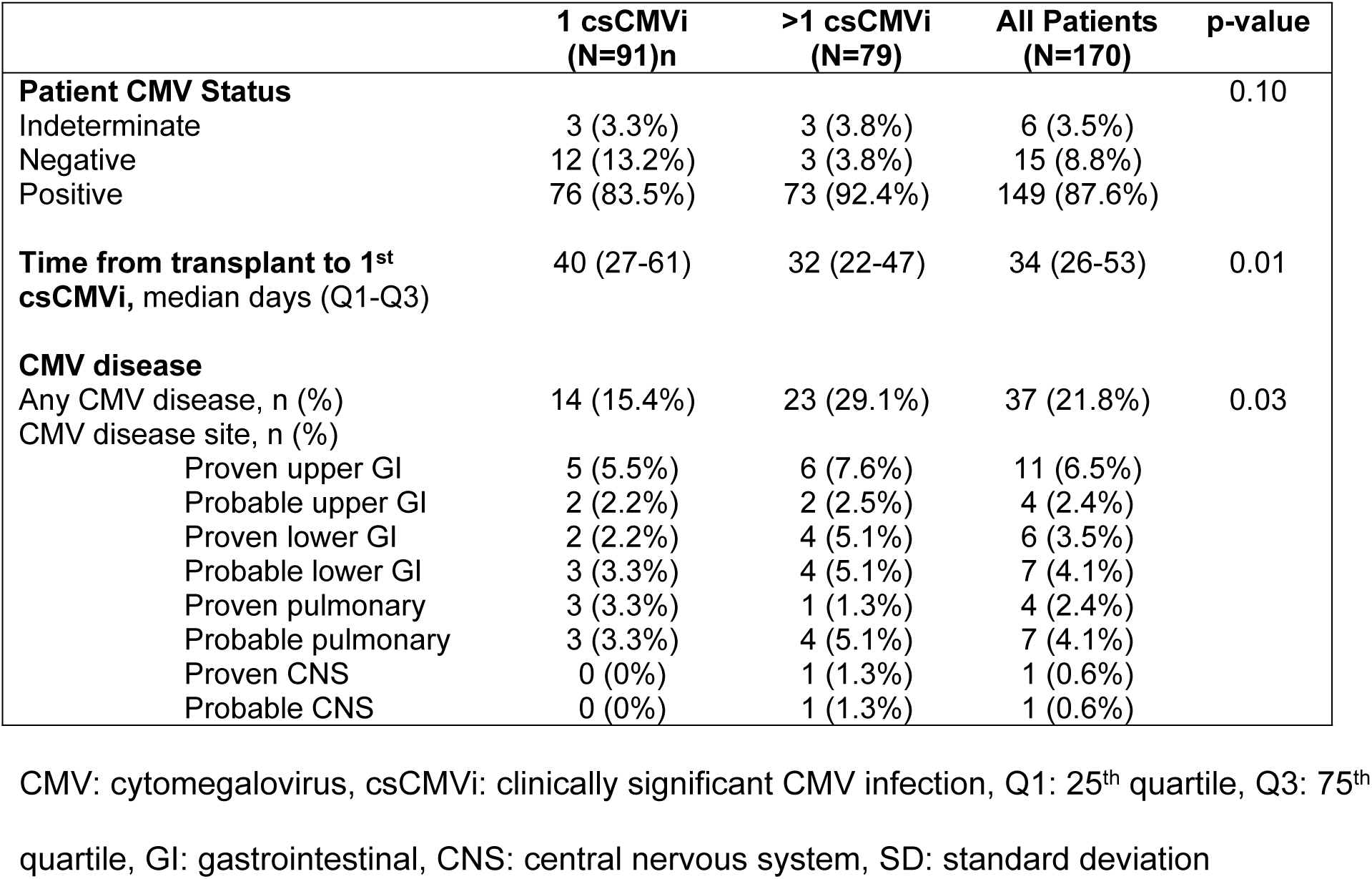
Clinically significant cytomegalovirus infection (csCMVi) characterization.

In comparison to patients with one episode of csCMVi, patients with multiple episodes of csCMVi were significantly more likely to have a transplant from an unrelated donor (72.2% versus 53.8%; p=0.014), have a cord blood transplant (26.6% vs. 12.1%; p=0.024), and to have received mycophenolate plus alemtuzumab for GVHD prophylaxis (31.1% vs. 19.8%; p=0.007) or mycophenolate plus a calcineurin inhibitor (33.8% vs. 18.6%; p=0.007) for GVHD prophylaxis (Table 1). Per Table 2, patients with multiple episodes of csCMVi had a shorter time from transplant to first csCMVi episode than patients with only one episode of csCMVi (median 32 days [Q1-Q3 22-47 days] vs. 40 days [22–47]; p<0.01) had significantly higher frequency of CMV disease (29.1% vs. 15.4%; p=0.03).

Antiviral resistance was more frequent among patients with multiple episodes of csCMVi compared to patients with one episode (6 patients, 7.6% versus 1 patient, 1.1%). For the single patient with resistant CMV infection during their first and only csCMVi, their csCMVi met the definition of probable refractory infection, prompting a switch from ganciclovir to foscarnet. The patient subsequently developed signs of CMV colitis, and CMV resistance evaluation revealed a UL54 gene mutation. Among the seven patients with resistant csCMVi, four patients had UL54 gene mutations, and three patients had UL97 mutations.

Few cases met the consensus definitions for refractory and probable refractory csCMVi and disease. We analyzed cases of refractory and probable refractory infections by CMV episode. Among patients with one episode of csCMVi, one episode met criteria for probable refractory csCMVi. Among multiple episodes of csCMVi, only three episodes met criteria for refractory csCMVi, eight episodes met criteria for probable refractory csCMVi, five episodes met criteria for refractory CMV disease, and one episode met criteria for probable refractory CMV disease.

Per Table 3, 59 (34.7%) of patients had at least one episode of bacteremia post-HCT, and the majority of these infections were monomicrobial (50 out of 59; 84.7%) and due to gram-positive organisms (31 out of 59; 52.5%). Twenty patients (11.8%) had a proven or probable IFD with the lung being the most common site of infection (14 out of 20; 70%). There was no significant difference in the rates of bacteremia or IFD based on multiple versus one episode of csCMVi.

**Table 3:** Comparison of bloodstream infections and invasive infections based on the number of clinically significant cytomegalovirus infection (csCMVi) episodes.

|  | 1 csCMVi<br>(N=91) | >1 csCMVi<br>(N=79) | All Patients<br>(N=170) | p value |
| --- | --- | --- | --- | --- |
| **Number of Bacteremia Episodes** |  |  |  | 0.80 |
| 1 episode | 26 (28.6%) | 24 (30.4%) | 50 (29.4%) |  |
| ≥ 2 episodes | 4 (4.4%) | 5 (6.3%) | 9 (5.3%) |  |
| **Bacteremia type** |  |  |  | 0.76 |
| Monomicrobial | 25 (83.3%) | 25 (86.2%) | 50 (84.7%) |  |
| Polymicrobial | 5 (16.7%) | 4 (13.8%) | 9 (15.3%) |  |
| **Gram-status Bacterium** |  |  |  | 0.92 |
| Gram-Positive | 15 (50.0%) | 16 (55.2%) | 31 (52.5%) |  |
| Gram-Negative | 14 (46.7%) | 12 (41.4%) | 26 (44.1%) |  |
| Nocardia/ Mycobacteria | 1 (3.3%) | 1 (3.4%) | 2 (3.4%) |  |
| **IFD Category<sup>a</sup>** |  |  |  | 0.20 |
| Probable | 3 (27.3%) | 5 (55.6%) | 8 (40.0%) |  |
| Proven | 8 (72.7%) | 4 (44.4%) | 12 (60.0%) |  |
| **IFD Type** |  |  |  | 0.78 |
| Aspergillosis | 2 (18.2%) | 4 (44.4%) | 6 (30.0%) |  |
| *Candida* | 1 (9.1%) | 1 (11.1%) | 2 (10.0%) |  |
| Mold, other | 1 (9.1%) | 0 (0.0%) | 1 (5.0%) |  |
| Mucormycosis | 4 (36.4%) | 2 (22.2%) | 6 (30.0%) |  |
| PJP | 2 (18.2%) | 1 (11.1%) | 3 (15.0%) |  |
| Unspecified IFD | 1 (9.1%) | 1 (11.1%) | 2 (10.0%) |  |
| **IFDSite** |  |  |  | 0.43 |
| Bloodstream | 1 (9.1%) | 1 (11.1%) | 2 (10.0%) |  |
| Disseminated | 2 (18.2%) | 0 (0.0%) | 2 (10.0%) |  |
| Lung | 7 (63.6%) | 7 (77.8%) | 14 (70.0%) |  |
| Rhinocerebral | 1 (9.1%) | 0 (0.0%) | 1 (5.0%) |  |
| Sinonasal | 0 (0.0%) | 1 (11.1%) | 1 (5.0%) |  |

IFD (invasive fungal disease), PJP (*Pneumocystis jirovecii* pneumonia)
a. Invasive fungal infection type defined by European Organization for Research and Treatment of Cancer and the Mycoses Study Group Education and Research Consortium [17].

Patients with multiple episodes of csCMVi were significantly more likely to receive treatment with foscarnet than patients with one episode of csCMVi (25.3% vs. 9.9%; p=0.01). The impact of csCMVi on renal function post-HCT was assessed by comparing changes in eGFR, incidence of AKI and acute renal failure at 90 days, 6 months, and 12 months post-transplant (Table 4). Patients with any csCMVi had significantly greater decline in eGFR at 12 months post-transplant compared with those never experiencing a csCMVi. Patients with any csCMVi also had significantly higher rates of AKI at 12 months post-transplant than patients without csCMVi (17.6% vs.11%; p=0.03). There was no difference in the rate of renal failure between the groups at any time point.

**Table 4.** Renal Function in patients with any clinically significant CMV infection (csCMVi) versus patients without clinically significant CMV infection.

| Characteristic | Any csCMVi<br>N=170 (43.8%) | No csCMVi<br>N=218 (56.2%) | All Patients<br>N=388 (100%) | p-value |
| --- | --- | --- | --- | --- |
| Change in eGFR 90 days post-transplant; Median (Q1-Q3) | -19 (-43.0 - 0) | -27 (-48.0 - 0) | -22.9 (-46.05 - 0) | 0.11 |
| Change in eGFR at 6 months | -21.22 (-42.8 - 0) | -25.79 (-47.4 - -6.88) | -23.64 (-44.87 - -5.66) | 0.18 |
| Change in eGFR at 12 months | -31.86 (-51.25 - -18.46) | -28.47 (-43.03 - -9.09) | -29.3 (-46.49 - -12.78) | 0.04 |
| eGFR decreased by 20% at 90 days | 76 (44.7%) | 106 (48.6%) | 182 (46.9%) | 0.09 |
| eGFR decreased by 30% at 90 days | 60 (35.3%) | 77 (35.3%) | 137 (35.3%) | 0.50 |
| eGFR decreased by 20% at 6 months | 78 (45.9%) | 93 (42.7%) | 171 (44.1%) | 0.45 |
| eGFR decreased by 30% at 6 months | 53 (31.2%) | 64 (29.4%) | 117 (30.2%) | 0.54 |
| eGFR decreased by 20% at 12 months | 64 (37.6%) | 72 (33%) | 136 (35.1%) | 0.11 |
| eGFR decreased by 30% at 12 months | 48 (28.2%) | 47 (21.6%) | 95 (24.5%) | 0.05 |
| Acute kidney failure at 100 days | 34 (20%) | 55 (25.2%) | 89 (22.9%) | 0.22 |
| Acute kidney injury at 100 days | 84 (49.4%) | 125 (57.3%) | 209 (53.9%) | 0.11 |
| Acute kidney injury at 6 months | 33 (19.4%) | 50 (22.9%) | 83 (21.4%) | 0.17 |
| Acute kidney failure at 6 months | 5 (2.9%) | 7 (3.2%) | 12 (3.1%) | 0.69 |
| Acute kidney injury at 12 months | 30 (17.6%) | 24 (11%) | 54 (13.9%) | 0.03 |
| Acute kidney failure at 12 months | 4 (2.4%) | 4 (1.8%) | 8 (2.1%) | 0.69 |
| Received foscarnet | 29 (17.1%) | 22 (10.1%) | 51 (13.1%) | 0.05 |

eGFR: estimated glomerular filtration rate expressed as mL/min/1.73 m<sup>2</sup>, Q1: 25<sup>th</sup> quartile, Q3: 75<sup>th</sup> quartile

When comparing patients with one episode of csCMVi to patients with multiple episodes of csCMVi, patients with multiple csCMVi had significantly larger declines in eGFR at 12 months (reduction by a median of -37.99mL/min/1.73m^2^ [-59.87 to -25.68] vs. -26.2 [-44.08 to -9.78]; p<0.01; Table 5). At 12 months post-transplant, a significant difference was observed in the percentage of patients with a 30% decrease in eGFR (36.7% vs. 20.9%; p=0.02).

**Table 5.** Comparison of renal function based on number of clinically significant cytomegalovirus infection (csCMVi) episodes.

|  | 1 csCMVi<br>N=91 (53.5%) | >1 csCMVi<br>N=79 (46.5%) | All Patients with<br>csCMVi<br>N=170 (100%) | p-value |
| --- | --- | --- | --- | --- |
| Change in eGFR at 90 days post-transplant; Median (Q1-Q3) | -20.63 (-41.35 - -7.44) | -18.47 (-45.03 - 0) | -19.13 (-42.74 - 0) | 0.64 |
| Change in eGFR at 6 months | -26.64 (-42.26 - -4.91) | -18.19 (-43.27 - 0) | -21.22 (-42.8 - 0) | 0.54 |
| Change in eGFR at 12 months | -26.2 (-44.08 - -9.78) | -37.99 (-59.87 - -25.68) | -31.86 (-51.25 - -18.46) | <0.01 |
| eGFR decreased by 20% 90 days | 39 (42.9%) | 37 (46.8%) | 76 (44.7%) | 0.75 |
| eGFR decreased by 30% at 90 days | 32 (35.2%) | 28 (35.4%) | 60 (35.3%) | 0.51 |
| eGFR decreased by 20% by at 6 months | 40 (44%) | 38 (48.1%) | 78 (45.9%) | 0.33 |
| eGFR decreased by 30% at 6 months | 27 (29.7%) | 26 (32.9%) | 53 (31.2%) | 0.53 |
| eGFR decreased by 20% at 12 months | 29 (31.9%) | 35 (44.3%) | 64 (37.6%) | 0.07 |
| eGFR decreased by 30% at 12 months | 19 (20.9%) | 29 (36.7%) | 48 (28.2%) | 0.02 |
| Acute kidney failure at 100 days | 20 (22%) | 14 (17.7%) | 34 (20%) | 0.49 |
| Acute kidney injury at 100 days | 50 (54.9%) | 34 (43%) | 84 (49.4%) | 0.12 |
| Acute kidney injury at 6 months | 19 (20.9%) | 14 (17.7%) | 33 (19.4%) | 0.22 |
| Acute kidney failure at 6 months | 1 (1.1%) | 4 (5.1%) | 5 (2.9%) | 0.20 |
| Acute kidney injury at 12 months | 13 (14.3%) | 17 (21.5%) | 30 (17.6%) | 0.29 |
| Acute kidney failure at 12 months | 1 (1.1%) | 3 (3.8%) | 4 (2.4%) | 0.28 |
| Received foscarnet | 9 (9.9%) | 20 (25.3%) | 29 (17.1%) | 0.01 |

eGFR: estimated glomerular filtration rate expressed as mL/min/1.73 m<sup>2</sup>, Q1: 25<sup>th</sup> quartile, Q3: 75<sup>th</sup> quartile

There were no significant differences in one- and five-year OS or NRM based on csCMVi versus no csCMVi, or number of episodes of csCMVi (Figures 1 and 2).

**Figure 1:**
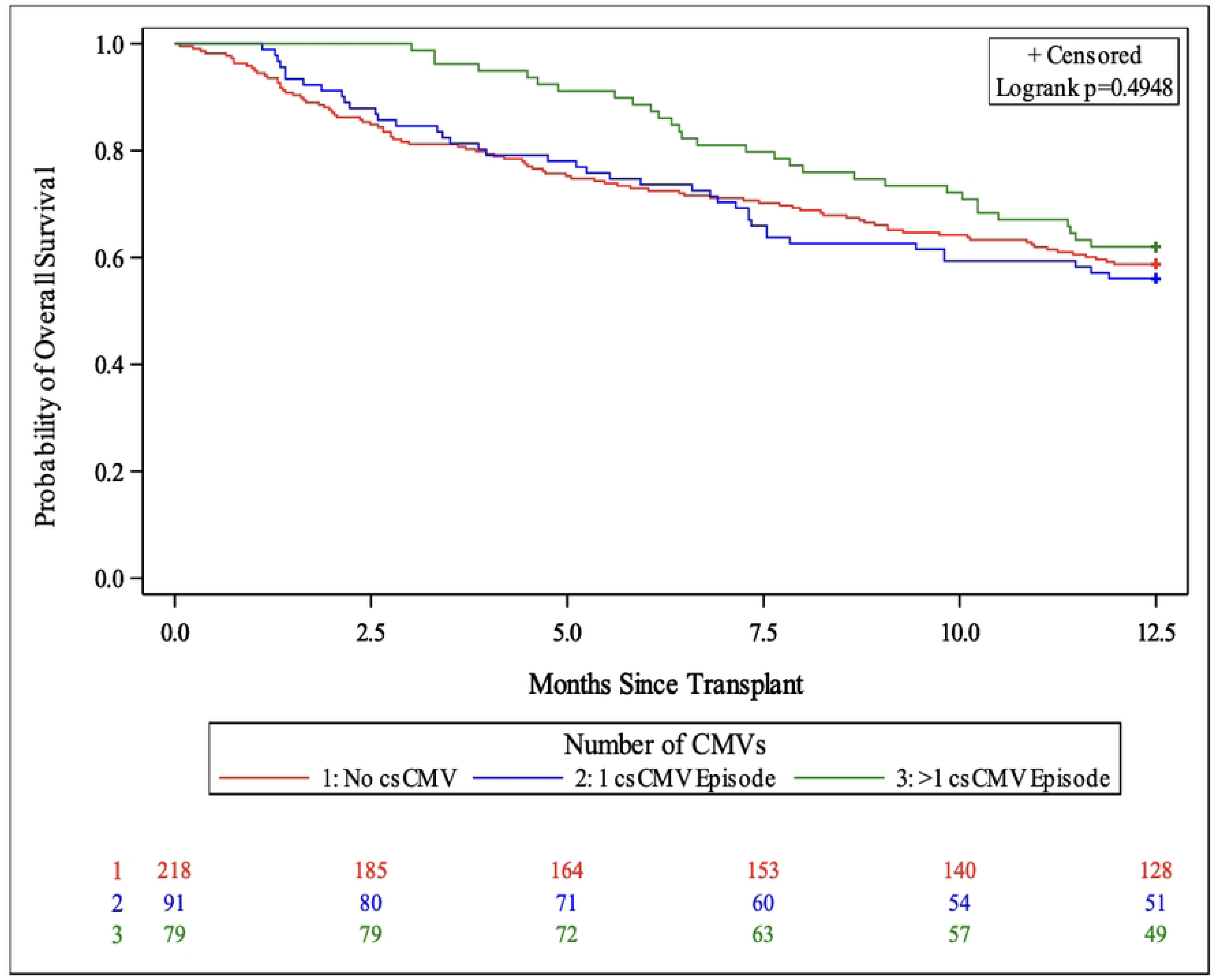
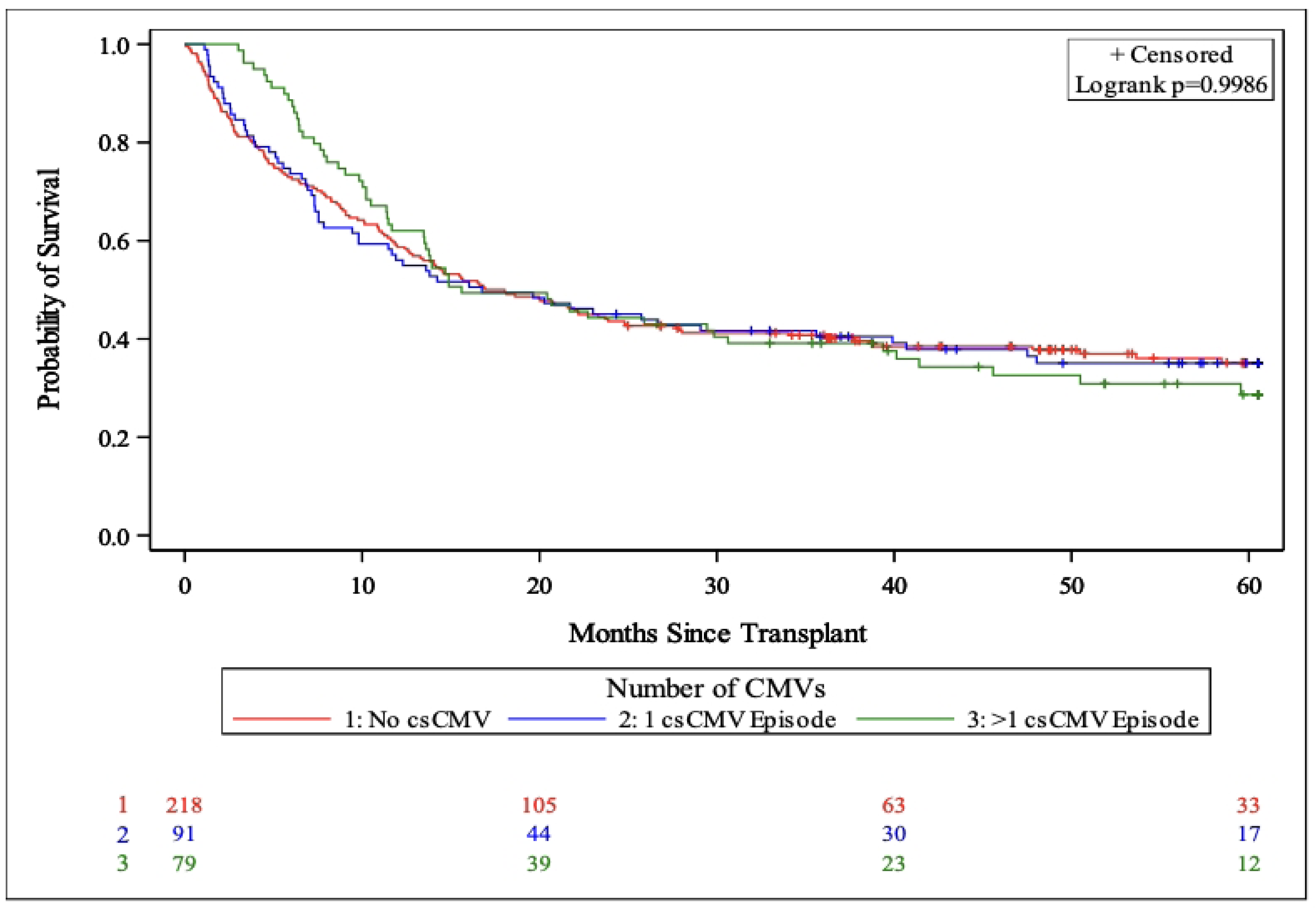
Overall survival for three groups: One csCMVi episode, greater than one csCMVi episode, and no csCMVi at one-year (A) and five-years post-transplant (B).

**Figure 2.**
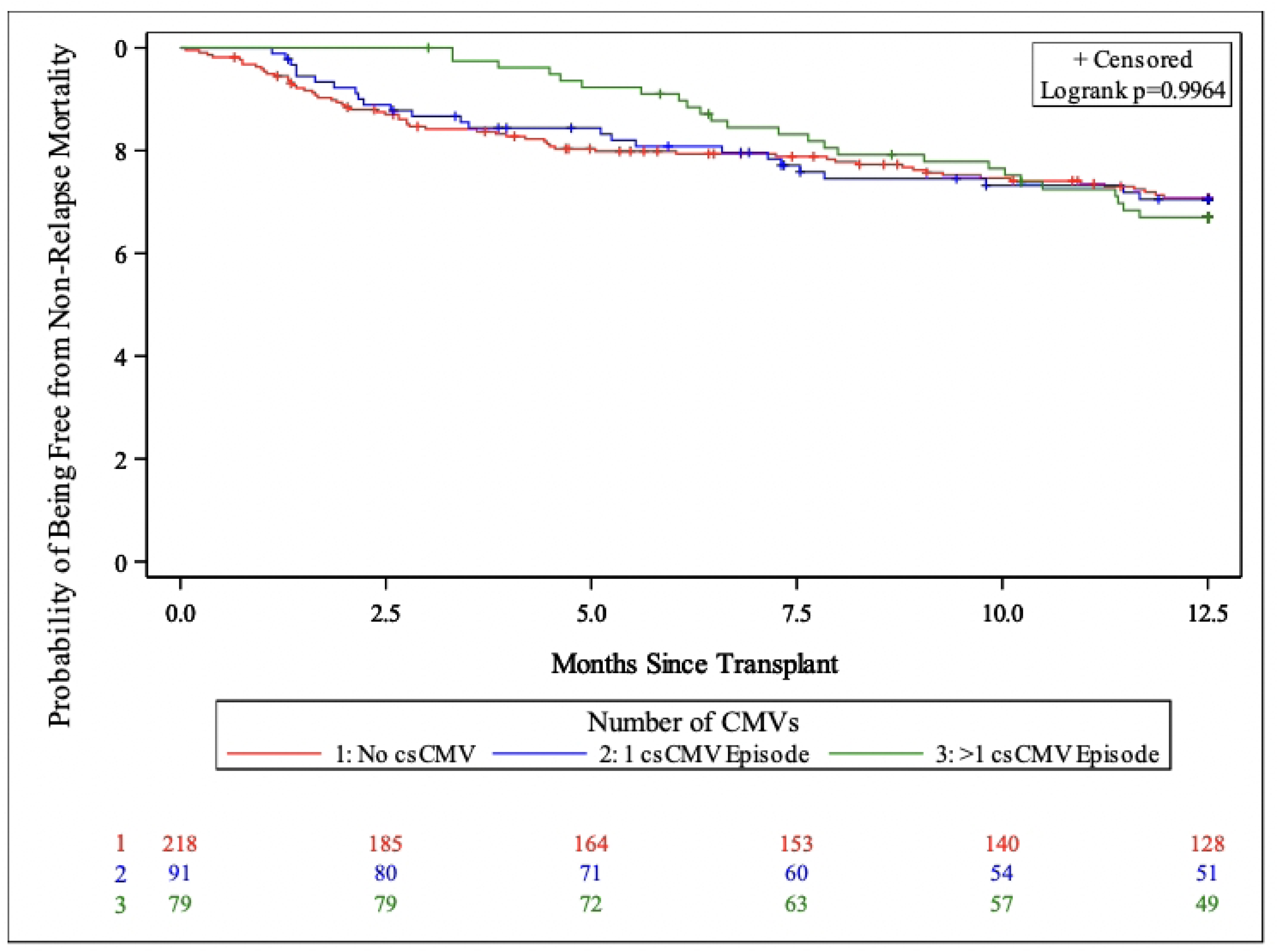
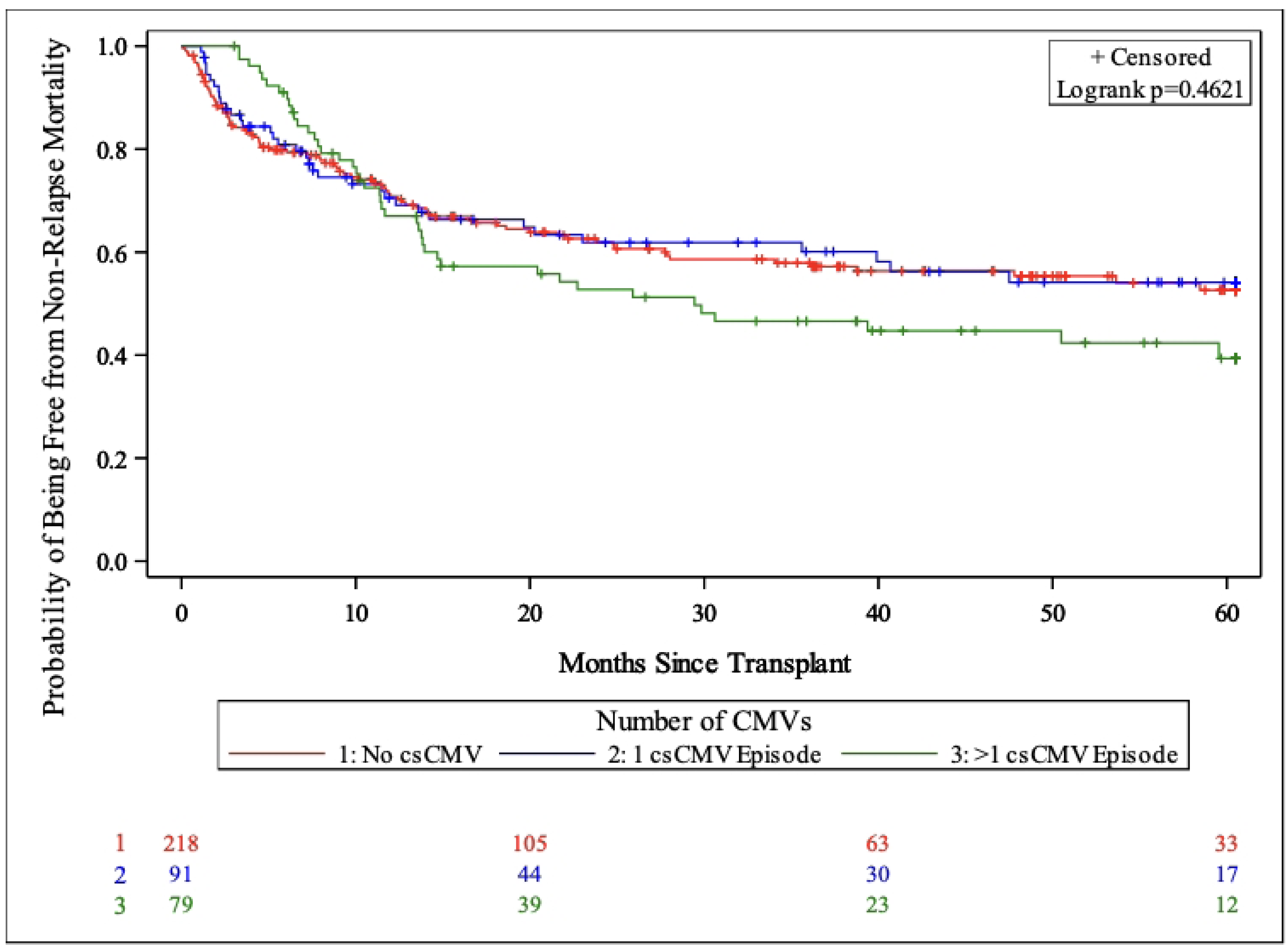
Non-relapse mortality (NRM) for three groups: one csCMVi episode, greater than one csCMVi episode, and no csCMVi at one-year (A) and (B) five-years post-transplant.

There was a significant decrease one-year RRM in patients with multiple episodes of csCMVi compared to patients with one episode of csCMVi or no csCMVi (p=0.04; Figure 3A). Additionally, we observed a trend toward increased incidence of five-year RRM in patients without csCMVi compared to patients with one episode of csCMVi or multiple episodes of csCMVi, but this comparison was not statistically significant (p=0.12; Figure 3B).

**Figure 3:**
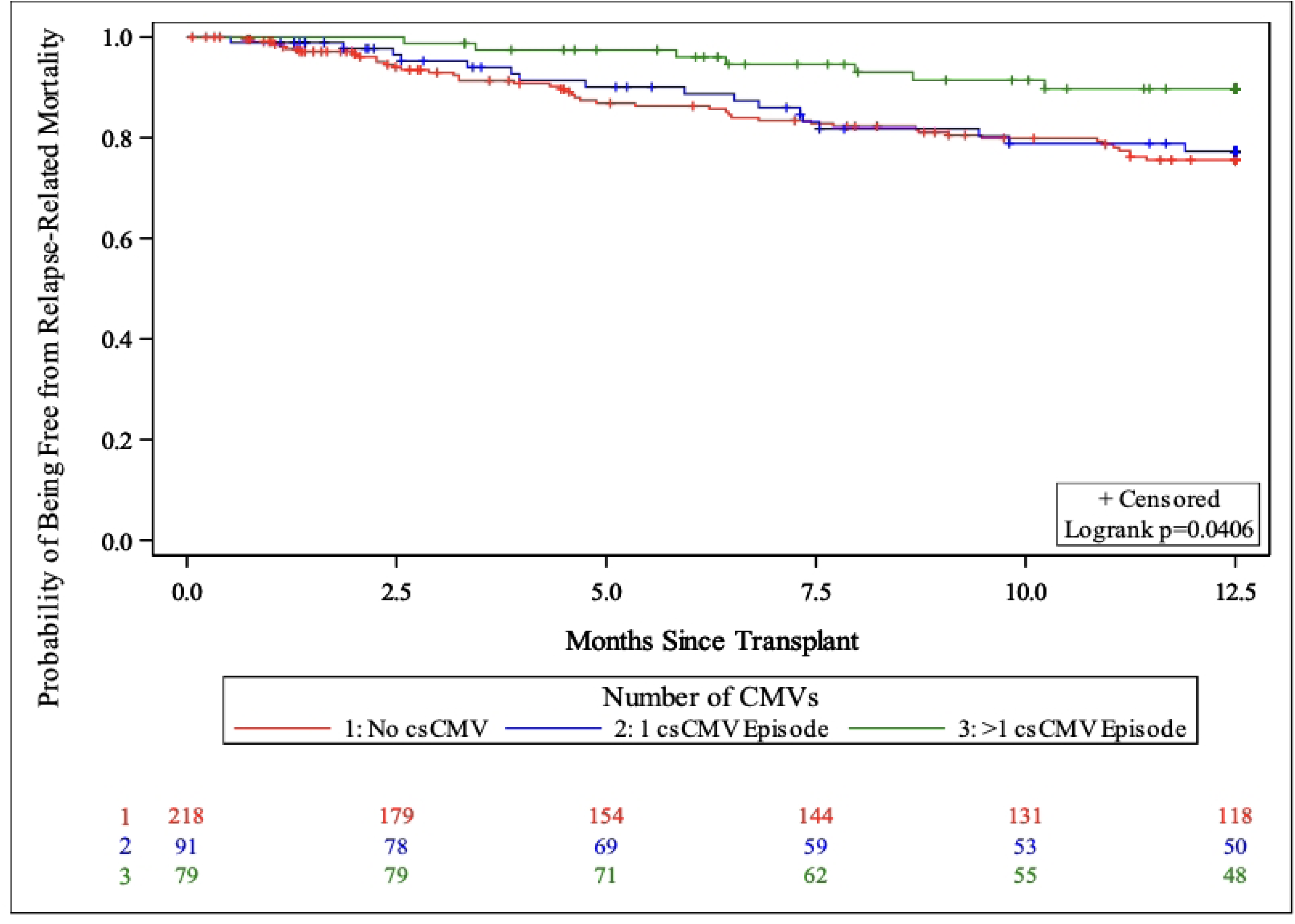
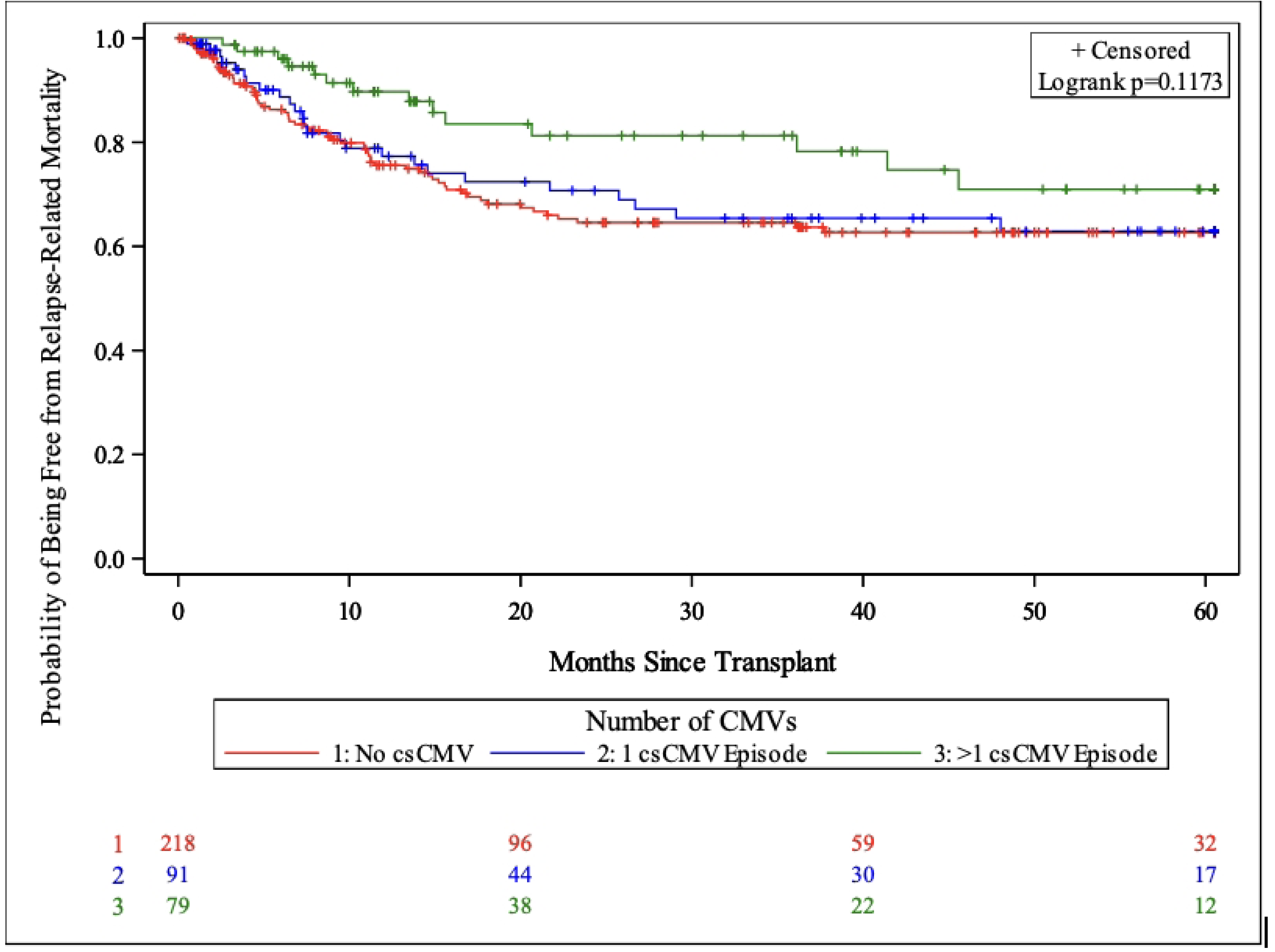
Disease relapse-related mortality (RRM) for three groups: one clinically significant CMV infection (csCMVi) episode, greater than one csCMVi episode, and no csCMVi at one-year (A) and five-years (B) post-transplant.

In the present study, we identify having a transplant from a matched unrelated donor, stem cell source as cord blood, and GVHD prophylaxis with highly T-cell suppressive agents, such as alemtuzumab, as risk factors for multiple episodes of csCMVi. Patients with multiple episodes tend to develop csCMVi earlier post-transplant and are more likely to have CMV disease. We did not find evidence to suggest patients with multiple episodes of csCMV had worse survival but do have lower one-year RRM compared to patients with one episode of csCMVi.

Prior reports of allogeneic HCT recipients experiencing multiple CMV episodes are limited to the early post-transplant time period. A study from Japan examined clinical outcomes and costs related to CMV within first 180 days post-transplant[18]. This retrospective study included patients who underwent transplant from April 2010 to March 2018 but did not differentiate whether patients received pre-emptive therapy versus universal prophylaxis. In thiss study, 64% of patients (479 out of 752 patients) had multiple episodes of CMV infection in comparison to 46% of patients (79 out of 170) in the present study.

In contrast to prior studies examining the impact of CMV infection on clinical outcomes, we did not observe a significant difference in bacteremia or IFD based on number of CMV episodes. This finding might have been impacted by our use of mold-active antifungal prophylaxis for at least the first 100 days post allogeneic HCT. Regarding prior relationships between csCMVi and IFD, in a multicenter retrospective cohort study in Australia, investigators found that CMV reactivation was an independent risk factor for IFD [2]. IFD occurred significantly later post-transplant in patients with CMV reactivation (median 184 days) compared to patients without CMV reactivation (37 days). Notably, having multiple episodes of CMV reactivation did not predict risk for IFD in the setting of standard antifungal prophylaxis.

Regarding renal dysfunction, our finding of more long-term kidney dysfunction is consistent with prior reports looking at the impact of foscarnet use on renal function in transplant recipients. One study examined the impact of foscarnet treatment for ganciclovir resistant or refractory CMV on outcomes among 39 solid organ transplant and HCT recipients[19]. In this study, renal dysfunction, defined as a >20% eGFR, was observed in 51% of patients between the start and end of foscarnet therapy and in 28% of the patients, renal dysfunction persisted six months after foscarnet therapy.

As was found in a prior study of foscarnet use in allogeneic HCT recipients, we observed a similar signal of long-term decline of renal function at the 12-month post-transplant time point in patients with multiple episodes of csCMVi [14]. Of note, both studies were performed at Duke University Medical Center during overlapping time periods (2009-13 for the present study and 2002-15 for the foscarnet study). In addition, we have demonstrated that patients with multiple episodes of csCMVi had higher frequencies of treatment with foscarnet. Regardless, the repeated observation of more long-term renal dysfunction in patients with multiple episodes of csCMVi and foscarnet exposure is important to keep in mind as a risk for chronic kidney disease post-allogeneic HCT.

In the present study, we did not observe an increase in non-relapse-related (treatment-related) mortality among patients with multiple episodes of csCMVi. In a prior study of the impacts of CMV serostatus on clinical outcomes in allogeneic HCT recipients, investigators determined CMV donor seropositive / recipient seronegative cases were at the highest risk for one-year mortality due to bacteremia or fungal infection [20]. However, when high CMV viral load and receipt of ganciclovir were introduced into the multivariable model, the association between CMV serostatus and mortality was not as pronounced.

In our analysis of mortality, patients with multiple episodes of csCMVi had significantly lower one-year relapse-related mortality. The impact of csCMVi on relapse-related mortality has not been studied as frequently as the impact on overall survival or non-relapse related mortality. A prior retrospective cohort study observed significantly lower risk of leukemia relapse among reduced-intensity allo HCT recipients of CMV-seropositive donors; however, this effect was offset by these same recipients having higher treatment-related mortality [21]. In another retrospective study, which was multicenter cohort of 9469 patients with leukemia and myelodysplastic disorder through the Center for International Blood and Bone Marrow Transplant Research, early CMV reactivation occurring prior to Day +100 was not associated with reduced risk for disease relapse but was associated with increased risk for NRM [2].

In the present study, we did not evaluate the occurrence of acute GVHD and compare rates of GVHD between patients with and without multiple episodes of csCMVi. One potential hypothesis as to why we observe reduced relapse-related mortality in patients with multiple episodes of csCMVi is that these patients also had more GVHD, which could infer more graft-versus-tumor effect to prevent relapse.

In the present study, we only identified nine csCMVi episodes that met criteria for probable refractory CMV infection, three episodes that met criteria for refractory CMV infection, one episode that met criteria for probable refractory CMV disease, and five episodes that met criteria for refractory CMV disease. Based on the low incidence of probable refractory or refractory csCMVi, we did not pursue further analysis of correlations with clinical outcomes. In contrast, a prior study of 488 allogeneic HCT recipients with CMV infection diagnosed with pre-emptive monitoring revealed that 50.6% of recipients met consensus criteria for refractory infection [1]. Risk factors for refractory infection included receipt of transplant from HLA-mismatched donors and development of acute GVHD. Recipients with refractory CMV infection had an increased incidence of CMV disease and higher NRM. In this prior study, there was no report of association of invasive infections and refractory CMV infection that may confound association with increased NRM.

Regarding CMV resistance, we did not see any cases of both UL97 and UL54 mutations. We did observe four cases of UL54 mutations (without UL96 mutations), which could be explained by our institution’s frequent use of foscarnet for prevention of complications from human herpes virus-6 reactivation or upfront treatment of CMV, over ganciclovir, due to cytopenias.

Strengths of the present study include clinical outcomes data for one-year post-transplant, focus on a population that underwent pre-emptive CMV monitoring, and the utilization of consensus definitions of csCMVi and refractory and resistant infection. We acknowledge multiple limitations of this study including its retrospective design taking place over a 5-year period from 2009-2013. We have intentionally chosen this time period as it reflects the pre-letermovir prophylaxis era for CMV high-risk HCT recipients. In the present study, we are reporting clinical outcomes associated with multiple episodes of csCMVi identified by pre-emptive monitoring to serve as a comparison group for future study of allogeneic HCT cohorts in the post-letermovir prophylaxis era.

## Conclusions

Patients with multiple episodes of csCMVi have higher frequency of CMV disease and although they have higher rates of antiviral resistance, rates of antiviral resistance and refractory CMV infection were surprisingly low in this cohort. In addition, we did not find an association between multiple episodes of csCMVi and rates of invasive bacterial and fungal infections. We did find that patients with multiple episodes of csCMVi had reduced one-year RRM compared to patients with one episode of csCMVi. Future study should focus on clinical outcomes of csCMVi and re-evaluate rates of multiple CMV episodes in cohorts that include patients receiving universal letermovir prophylaxis.

## Data Availability

All relevant data are within the manuscript and its Supporting Information files.

